# Genome-wide association meta-analysis of childhood and adolescent internalising symptoms

**DOI:** 10.1101/2020.09.11.20175026

**Authors:** Eshim S Jami, Anke R Hammerschlag, Hill F Ip, Andrea G Allegrini, Beben Benyamin, Richard Border, Elizabeth W Diemer, Chang Jiang, Ville Karhunen, Yi Lu, Qing Lu, Travis T Mallard, Pashupati P Mishra, Ilja M Nolte, Teemu Palviainen, Roseann E Peterson, Hannah M Sallis, Andrey A Shabalin, Ashley E Tate, Elisabeth Thiering, Natàlia Vilor-Tejedor, Carol Wang, Ang Zhou, Daniel E Adkins, Silvia Alemany, Helga Ask, Qi Chen, Robin P Corley, Erik A Ehli, Luke M Evans, Alexandra Havdahl, Fiona A Hagenbeek, Christian Hakulinen, Anjali K Henders, Jouke Jan Hottenga, Tellervo Korhonen, Abdullah Mamun, Shelby Marrington, Alexander Neumann, Kaili Rimfeld, Fernando Rivadeneira, Judy L Silberg, Catharina E van Beijsterveldt, Eero Vuoksimaa, Alyce M Whipp, Tong Xiaoran, Ole A Andreassen, Dorret Boomsma, Sandra A Brown, S Alexandra Burt, William Copeland, Elizabeth J Costello, Danielle M Dick, Lindon J Eaves, K Paige Harden, Kathleen Mullan Harris, Catharina A Hartman, Joachim Heinrich, John K Hewitt, Christian Hopfer, Elina Hypponen, Marjo-Riitta Jarvelin, Jaakko Kaprio, Liisa Keltikangas-Järvinen, Kelly L Klump, Kenneth Krauter, Ralf Kuja-Halkola, Henrik Larsson, Terho Lehtimäki, Paul Lichtenstein, Sebastian Lundstrom, Hermine H Maes, Per Magnus, Marcus R Munafò, Jake M Najman, Pål R Njølstad, Albertine J Oldehinkel, Craig E Pennell, Robert Plomin, Ted Reichborn-Kjennerud, Chandra Reynolds, Richard J Rose, Andrew Smolen, Harold Snieder, Michael Stallings, Marie Standl, Jordi Sunyer, Henning Tiemeier, Sally Wadsworth, Tamara L Wall, Andrew J O Whitehouse, Gail M Williams, Eivind Ystrom, Michel G Nivard, Meike Bartels, Christel M Middeldorp

**Affiliations:** Department of Biological Psychology, Vrije Univsiteit Amsterdam, Amsterdam, the Netherlands; Department of Clinical, Educational and Health Psychology, Division of Psychology and Language Sciences, University College London, London, UK; Amsterdam Public Health Research Institute, Amsterdam, the Netherlands; Child Health Research Centre, University of Queensland, Brisbane, Australia; Social, Genetic and Developmental Psychiatry Centre, King’s College London, London, UK; Australian Centre for Precision Health, University of South Australia Cancer Research Institute, Adelaide, Australia; South Australian Health and Medical Research Institute, Adelaide, Australia; Institute for Behavioral Genetics, University of Colorado Boulder, Boulder, CO, USA; Department of Child Psychiatry, Erasmus University Medical Center, Rotterdam, the Netherlands; Department of Psychology, Michigan State University, East Lansing, USA; Department of Biostatistics, University of Florida, Gainesville, USA; Department of Epidemiology and Biostatistics, Imperial College London, London, UK; Department of Medical Epidemiology and Biostatics, Karolinska Institutet, Stockholm, Sweden; Department of Epidemiology and Biostatistics, Michigan State University, East Lansing, USA; Department of Psychology, University of Texas, Austin, Texas, USA; Department of Clinical Chemistry, Fimlab Laboratories, and Finnish Cardiovascular Research Center, Tampere University, Tampere, Finland; Department of Epidemiology, University of Groningen, University Medical Center Groningen, Groningen, the Netherlands; Institute for Molecular Medicine Finland - FIMM, University of Helsinki, Helsinki, Finland; Virginia Institute for Psychiatric and Behavioral Genetics, Department of Human & Molecular Genetics, Virginia Commonwealth University, Richmond, VA, USA; School of Psychological Science, University of Bristol, Bristol, UK; MRC Integrative Unit, University of Bristol, Bristol, UK; Centre for Academic Mental Health, Population Health Sciences, University of Bristol, Bristol, UK; Department of Psychiatry, University of Utah, Salt Lake City, UT, USA; Institute of Epidemiology, Helmholtz Zentrum München - German Research Center for Environmental Health, Neuherberg, Germany; LMU – Ludwig-Maximilians-Universität Munich, Dr. von Hauner Children’s Hospital, University of Munich Medical Center, Munich, Germany; Computational Biology of RNA Processing, Centre for Genomic Regulation (CRG),The Barcelona Institute of Science and Technology, Barcelona, Spain; BarcelonaBeta Brain Research Center, (BBRC). Pasqual Maragall Foundation, Barcelona, Spain; Department of Clinical Genetics, Erasmus Medical Center, Rotterdam, the Netherlands; Universitat Pompeu Fabra (UPF), Barcelona, Spain; School of Medicine and Public Health, Faculty of Medicine and Health, University of Newcastle, Newcastle, NSW, Australia; Departments of Psychiatry and Sociology, University of Utah, Salt Lake City, UT, USA; Child and Environment Programme, ISGlobal, Barcelona Institute of Global Health, Barcelona, Spain; CIBER Epidemiología y Salud Pública (CIBERESP), Spain; Department of Mental Disorders, Norwegian Institute of Public Health, Oslo, Norway; Avera Institute for Human Genetics, Avera McKennan Hospital & University Health Center, Sioux Falls, USA; Institute for Behavioral Genetics, Department of Ecology & Evolutionary Biology, University of Colorado Boulder, Boulder, CO, USA; Department of Psychology and Logopedics, Faculty of Medicine, University of Helsinki, Helsinki, Finland; Institute for Molecular Biosciences, University of Queensland, Brisbane, Australia; Institute for Social Science Research, University of Queensland, Brisbane, Australia; School of Public Health, Faculty of Medicine, University of Queensland, Brisbane, Australia; Department of Child and Adolescent Psychiatry/Psychology, Erasmus University Medical Center, Rotterdam, the Netherlands; Lady Davis Institute for Medical Research, Jewish General Hospital, Montreal, Canada; The Generation R Study Group, Erasmus University Medical Center, Rotterdam, the Netherlands; Department of Epidemiology, Erasmus University Medical Center, Rotterdam, the Netherlands; Department of Internal Medicine, Erasmus University Medical Center, Rotterdam, the Netherlands; NORMENT Centre, Institute of Clinical Medicine, University of Oslo, Oslo, Norway; Division of Mental Health and Addiction, Oslo University Hospital, Oslo, Norway; Departments of Psychology and Psychiatry, University of California San Diego, La Jolla, CA, USA; Vermont Center for for Children, Youth and Familes in the Department of Psychiatry, University of Vermont, Burlington, VT, USA; Department of Psychiatry, Duke University School of Medicine, Durham, NC, USA; Departments of Psychology and Human and Molecular Genetics, Virginia Commonwealth University, Richmond, VA, USA; Department of Sociology, Carolina Population Center, University of North Carolina at Chapel Hill, Chapel Hill, NC, USA; Department of Psychiatry, Interdisciplinary Center Psychopathology and Emotion Regulation, University of Groningen, University Medical Center Groningen, Groningen, the Netherlands; Institute and Outpatient Clinic for Occupational, Social and Environmental Medicine, University Hospital of Ludwig-Maximilians-Universität, Munich, Germany; Allergy and Lung Health Unit, Melbourne School of Population and Global Health, The University of Melbourne, Victoria, Australia; Department of Psychiatry, University of Colorado, Aurora, USA; Department of Molecular, Cellular and Developmental Biology, University of Colorado Boulder, Boulder, CO, USA; Department of Medical Epidemiology and Biostatics and School of medical sciences, Karolinska Institutet, Stockholm and Örebro, Sweden; Gillberg Neuropsychiatry Centre, Centre of Ethics, Law and Mental Health, Institute of Neurscience and Physiology, University of Gothenburg, Gothenburg, Sweden; Centre for Fertility and Health, Norwegian Institute of Public Health, Oslo, Norway; NIHR Biomedical Research Centre at the University Hospitals Bristol NHS Foundation Trust and the University of Bristol, Bristol, UK; Center for Diabetes Research, Department of Clinical Science, Univerity of Bergen, Bergen, Norway; Department of Pediatrics and Adolescents, Haukeland University Hospital, Bergen, Norway; Department of Psychology, University of California at Riverside, Riverside, CA, USA; Department of Psychological & Brain Sciences, Indiana University, Bloomington, Indiana, USA; Child and Environment Programme, Barcelona Institute of Global Health, Barcelona, Spain; IMIM (Hospital del Mar Medical Research Institute), Barcelona, Spain; Department of Social and Behavioral Science, Harvard TH Chan School of Public Health, Boston, MA, USA; Department of Psychiatry, University of California, San Diego, La Jolla, CA, USA; Telethon Kids Institute, University of Western Australia, Perth, Australia; Department of Psychology, University of Oslo, Oslo, Norway; Child and Youth Mental Health Service, Children’s Health Queensland Hospital and Health Service, Brisbane, Australia

## Abstract

Internalising symptoms in childhood and adolescence are as heritable as adult depression and anxiety, yet little is known of their molecular basis. This genome-wide association meta-analysis of internalising symptoms included repeated observations from 64,641 individuals, aged between 3 and 18. The N-weighted meta-analysis of overall internalising symptoms (INT_overall_) detected no genome-wide significant hits and showed low SNP heritability (1.66%, 95% confidence intervals 0.84-2.48%, N_effective_=132,260). Stratified analyses indicated rater-based heterogeneity in genetic effects, with self-reported internalising symptoms showing the highest heritability (5.63%, 95% confidence intervals 3.08-8.18%). Additive genetic effects on internalising symptoms appeared stable over age, with overlapping estimates of SNP heritability from early-childhood to adolescence. Genetic correlations were observed with adult anxiety, depression, and the wellbeing spectrum (|*r*_*g*_|> 0.70), as well as with insomnia, loneliness, attention-deficit hyperactivity disorder, autism, and childhood aggression (range |*r*_*g*_|=0.42-0.60), whereas there were no robust associations with schizophrenia, bipolar disorder, obsessive-compulsive disorder, or anorexia nervosa. The pattern of genetic correlations suggests that childhood and adolescent internalising symptoms share substantial genetic vulnerabilities with adult internalising disorders and other childhood psychiatric traits, which could partially explain both the persistence of internalising symptoms over time and the high comorbidity amongst childhood psychiatric traits. Reducing phenotypic heterogeneity in childhood samples will be key in paving the way to future GWAS success.

## Introduction

Internalising disorders, including anxiety and depression, are substantial contributors to the global burden of disease (1, 2). Whilst the estimated 12-month prevalence of depression and anxiety disorders in adults is 15% (3), internalising disorders are also present in early life, with an estimated prevalence of 2-3% of depression and 6-7% of anxiety in childhood and adolescence (4). Prior to the diagnosis of internalising disorders, as many as one in five children self-report internalising symptoms (5). These early symptoms of anxiety and depression appear to pose a long-term risk, as longitudinal studies show that internalising symptoms in childhood are associated with mood disorders, anxiety, and suicidality in adulthood (6-8). Findings from twin research show that internalising symptoms have a moderately strong genetic component. 40-50% of individual differences in internalising symptoms are explained by genetic factors (9-11). Moreover, research suggests that both stability and change in anxious and depressive symptoms from early childhood to adulthood are genetically influenced (10, 12-14). However, unlike adult anxiety and depression, investigation of the molecular genetic architecture of internalising symptoms in early life has received little attention thus far and to date, only two studies have applied a genome-wide approach (15, 16).

Published in 2013 and 2014, the first genome-wide association studies (GWASs) on childhood internalising symptoms did not identify any genome-wide significant hits for maternal-reported anxiety-related behaviours in children aged seven (N=2,810) (15), or internalising problems in children aged three (N=4,596) (16). Estimates of SNP-based heritability (the proportion of phenotypic variance captured by single nucleotide polymorphisms (SNPs) included in the GWAS), using genome-wide complex trait analysis (GCTA), were not robust in both studies (15, 16). Other GCTA studies similarly show mostly inconsistent and broad estimates of SNP heritability, mainly due to small sample sizes (17-22). Large-scale GWASs have led to significant discoveries in adult samples, with now 102 variants identified for depression (23) and 5 variants for anxiety (24). Given the comparable heritability estimates of adult and childhood internalising phenotypes, the next step in this line of research is to increase childhood sample sizes in order to generate sufficient power to capture the small effects of common variants that have been observed in adult studies.

Here, we present a genome-wide association meta-analysis which aims to identify common genetic variants associated with the development and course of internalising symptoms. The study combines repeated measurements of dimensional symptom scores from 22 independent cohorts of European ancestry, resulting in an overall sample of 64,641 individuals and 251,152 observations in children and adolescents aged between 3 and 18. All datasets were combined to produce a GWAS of overall internalising symptoms (INT_overall_), with an effective sample size of 132,260. Stratified analyses were used to investigate age, rater, and instrument-specific genetic effects. The overall GWAS of INT_overall_ was followed up with gene-based analyses. Genetic overlap with external traits was examined by computing genetic correlations, with a focus on psychiatric phenotypes. Non-psychiatric traits were also investigated if they were previously found to be genetically correlated with adult anxiety and depression (23-25). Finally, polygenic scores were computed to test prediction of internalising symptoms in independent samples. With this study, we aim to gain insight into the genetic underpinnings of internalising symptoms throughout childhood and adolescence in order to improve our understanding of the development and progression of internalising disorders.

## Methods

This project was pre-registered at the Open Science Framework (https://osf.io/edas6). Minor deviations from the pre-registration are explained in the Supplementary Note.

### Sample and univariate analyses

The sample includes cohorts that are part of the EArly Genetics and Lifecourse Epidemiology (EAGLE) consortium behaviour and cognition working group (https://www.eagle-consortium.org/) (26) and additional cohorts with appropriate data. In total, 22 cohorts of European ancestry participated in the study. Ethical approval was provided by local committees at cohort level. Many cohorts were longitudinal birth or childhood cohort studies with long-term follow-up and multiple raters, e.g., mother, father, self and teacher. Repeated assessments of internalising symptoms within childhood and adolescence, from age 3 to age 18, were included. All cohorts performed univariate GWASs stratified by (i) age, (ii) rater, and (iii) instrument, with a minimum of 450 observations in each analysis. In the absence of diagnostic data, Internalising symptoms were dimensionally measured and positively scored on continuous scales, with higher scores indicating more internalising symptoms. Data was not dichotomized into a case-control design as this would have resulted in a reduction of statistical power (27). Detailed descriptions of the cohorts, phenotypic measures, and genotyping and imputation procedures can be found in Supplementary Tables 1-6 and the Supplementary Note.

Cohorts that included only unrelated subjects applied a linear regression model. Cohorts with a sample of related individuals corrected for non-independence of observations by either applying a mixed linear model or a sandwich correction of the standard errors. As there is no evidence for sex differences in genetic variation in psychiatric traits (28), analyses were not stratified by sex but gender was included as a covariate. Further details about the univariate GWASs are provided in the Supplementary Note. In total, 125 univariate GWASs were collated, with 251,152 observations based on 64,641 unique participants. The observations included ratings by mothers (40.7%), fathers (6.8%), teachers (18.3%), self (19.7%) and siblings (0.7%). An additional 13.8% of ratings were parental reports, where the informant was either the mother or the father. 15.1% of observations were in early-childhood (3 to 6 years), 36.0% in mid-childhood (7 to 10 years), 18.4% in late-childhood (11 to 12 years), and 30.0% in adolescence (13 to 18 years). Twelve instruments were used to measure internalising symptoms, of which the most commonly used were the Strengths and Difficulties Questionnaire (SDQ; 38.2%) (29), Achenbach System of Empirically Based Assessment (ASEBA; 36.7%) (30) and Rutter Children’s Questionnaires (8.2%) (31, 32).

### Meta-analyses and the calculation of SNP heritabilities stratified by age, rater, and instrument

Quality control for each univariate GWAS was performed using EasyQC (Supplementary Text) (33). After QC, most cohorts retained between 3.4 and 7.1 million autosomal SNPs per GWAS (Supplementary Table 7). An exception was the Philadelphia Neurodevelopmental Cohort which retained fewer SNPs after merging data from different genotyping platforms. To account for dependency of repeated measurements of internalising symptoms within cohorts, the N-weighed meta-analysis approach was applied (34, 35). In short, two N × N matrices, representing sample overlap and phenotypic covariance within cohorts, were created, where N was the total number of univariate GWASs. As there was no overlap across cohorts, sample overlap and phenotypic covariance between cohorts were set to zero. Using the observed sample overlap within cohorts and their phenotypic covariance matrices, expected pairwise cross-trait intercept (CTI) values between GWASs were calculated. The pairwise CTI is approximately equal to the covariance between the test statistics from univariate GWASs. N-weighted meta-analyses were performed to obtain a multivariate test statistic per SNP, which represents a weighted sum of test statistics, adjusted by the CTI in order to account for sample overlap between GWASs. Formulas for the calculation of the multivariate test statistic for each SNP in the meta-analyses, the CTI between GWASs and estimation of effective sample size to account for repeated measurements (N_eff_) are provided in Ip *et al*. supplementary text (35).

A meta-analysis was performed based on the results of all available GWASs on internalising symptoms: INT_overall_ SNP-based heritabilities (*h*^*2*^) were estimated using linkage disequilibrium score regression (LDSC) (36), first for INT_overall_, and next based on results of meta-analyses stratified according to rater, age, rater-by-age, and instrument (Supplementary Table 8). To ensure that the stratified analyses had sufficient power, a sample size threshold was set so that the total number of observations (N_obs_) for each meta-analysis was at least 15,000. Rater-specific SNP heritabilities were estimated using assessments from parents (mother and/or father), mothers, fathers, teachers, and self, respectively. Age-specific SNP heritabilities focused on internalising symptoms during early childhood (3 to 6 years), mid-childhood (7 to 10 years), late-childhood (11 to 12 years), and adolescence (13 to 18 years). Rater-by-age SNP heritabilities assessed age effects within and between raters, provided that the univariate N_obs_ exceeded 15,000. Lastly, instrument-specific SNP heritabilities were calculated for SDQ, ASEBA, and Rutter for which the N_obs_ exceeded 15,000.

Genetic correlations across stratified GWAMAs were calculated using LDSC, but only if the z-score of the heritability estimate was ≥ 4, given that the heritability z-score is a good indicator of power and a score less than 4 is considered too noisy for meaningful estimates (37).

SNPs with minor allele frequency < 5% or N_eff_ < 15,000 were removed from further analyses.

### Gene-based analysis

Using summary statistics for INT_overall_, a MAGMA (38) gene-based test (v1.8, implemented in FUMA (39)) was performed to identify genes with a significant effect on internalising symptoms. The gene-based test applies a multiple regression model in which *p*-values from individual SNPs in a gene are combined into a test-statistic for each gene, while accounting for linkage disequilibrium between SNPs. European populations from the 1000 Genomes Phase III reference panel were used to estimate linkage disequilibrium. A total of 18,592 protein-coding genes were assessed for an association with internalising symptoms. A Bonferroni correction was applied to correct for multiple testing (*α* = 0.05 / 18,592; *p* < 2.69×10^−06^).

### Tissue expression and gene-set analyses

Tissue enrichment and gene-set analyses were conducted in FUMA. The tissue enrichment analyses used two types of tissues from GTEx version 8: 30 general tissue types from multiple organs and 53 specific tissue types within these organs. A MAGMA gene-property test was performed to test one-sided relationships between cell type-specific gene expression and disease–gene associations. Bonferroni corrections were applied to correct for multiple testing for the general (*α* = 0.05 / 30; *p* < 1.7×10^−04^) and specific (*α* = 0.05 / 53; *p* < 9.4×10^−04^) tissue types.

The gene-set analysis was performed with default parameters in MAGMA v1.8. Gene-based P-values were converted to Z values and a between-gene correlation matrix was used as input to perform gene-set enrichment tests. Predefined gene sets from the molecular signature database MsigDB v7.0 were used. In total, 15,484 gene sets were tested. A Bonferroni correction was applied to correct for multiple testing (*α* = 0.05 / 15,484; *p* < 3.2×10^−06^).

### Genetic correlations with external traits

Genetic correlations between internalising symptoms and other phenotypes were investigated using publicly available summary statistics for a curated set of traits (N=27). These primarily included adult psychiatric traits, in addition to other phenotypes selected based on previously identified correlations with adult anxiety and depression (23-25). Additionally, we obtained summary statistics from the GWA meta-analyses of overall and rater-specific childhood and adolescent aggression (35), that were based on overlapping cohorts and similar statistical methods, and calculated genetic correlations with these traits. The external traits and source studies are summarised in Supplementary Table 9. Summary statistics from INT_overall_ and INT_self_ (for which the z-score of the *h*^*2*^ was ≥ 4 (37)) were used. Genetic correlations were calculated using LDSC (36), which calculates genetic covariance between two traits based on all polygenic effects captured by included SNPs. Overlapping samples or population differences in GWAS summary statistics do not bias the computation of genetic correlations in LDSC. LDSC corrects for sample overlap by including a covariance matrix of the cross-trait LD score intercept, which is an estimate of sample overlap and phenotypic correlation. The genetic correlation estimate was based on the estimated slope from regressing the product of z-scores from two GWASs on the LD score. The LD scores used were computed using 1000 Genomes Phase III European data (37). Genetic correlations were considered significant at *p* < 9.26×10^−04^, after applying a Bonferroni correction for 54 independent tests.

### Sensitivity analysis: polygenic score prediction

Polygenic score prediction of INT_overall_ was tested as a sensitivity analysis. The Netherlands Twin Register (NTR) was used as the target sample to examine prediction of internalising symptoms in childhood and adolescence. We considered maternal-reported internalising symptoms at age 7 (N=3,845), and self-reported internalising symptoms during adolescence (age 13 to 18, N=2,679), using the ASEBA Child Behaviour Checklist and the Youth Self Report scales (30), respectively. A leave-one-cohort-out meta-analysis omitting data from NTR was performed for INT_overall_. The NTR target dataset was restricted to SNPs with minor allele frequency > 5% and imputation quality of *R*^*2*^ > 90%. Polygenic scores were constructed using LDpred (40), using a prior value of 0.5 to account for high polygenicity. Associations between polygenic scores of internalising symptoms and internalising problems were examined using Generalized Estimating Equations as implemented in the “gee” package in R (version 3.5.2). To account for relatedness in the target sample, the exchangeable working correlation matrix in gee was used, which applies a sandwich correction over the standard errors to account for clustering in the data. Age, sex, genotyping array, and the first 10 genetic principal components were included as covariates. Polygenic prediction was considered significant at *p* < 0.025, after applying a Bonferroni correction for 2 independent tests.

## Results

### Overall meta-analysis of childhood and adolescent internalising symptoms

The genome-wide association meta-analysis of INT_overall_ found no genome-wide significant hits (Figure 1). Assuming a N_eff_ of 132,260, SNP-based heritability of INT_overall_ was estimated at 1.66% (95% confidence interval (CI) 0.84-2.48%). The mean chi-squared statistic was 1.086, with an LDSC-intercept of 1.043 (standard error (SE)= 0.0075), indicating that a small part of the inflation in test statistics might have been due to confounding biases, such as population stratification.

**Figure 1.**
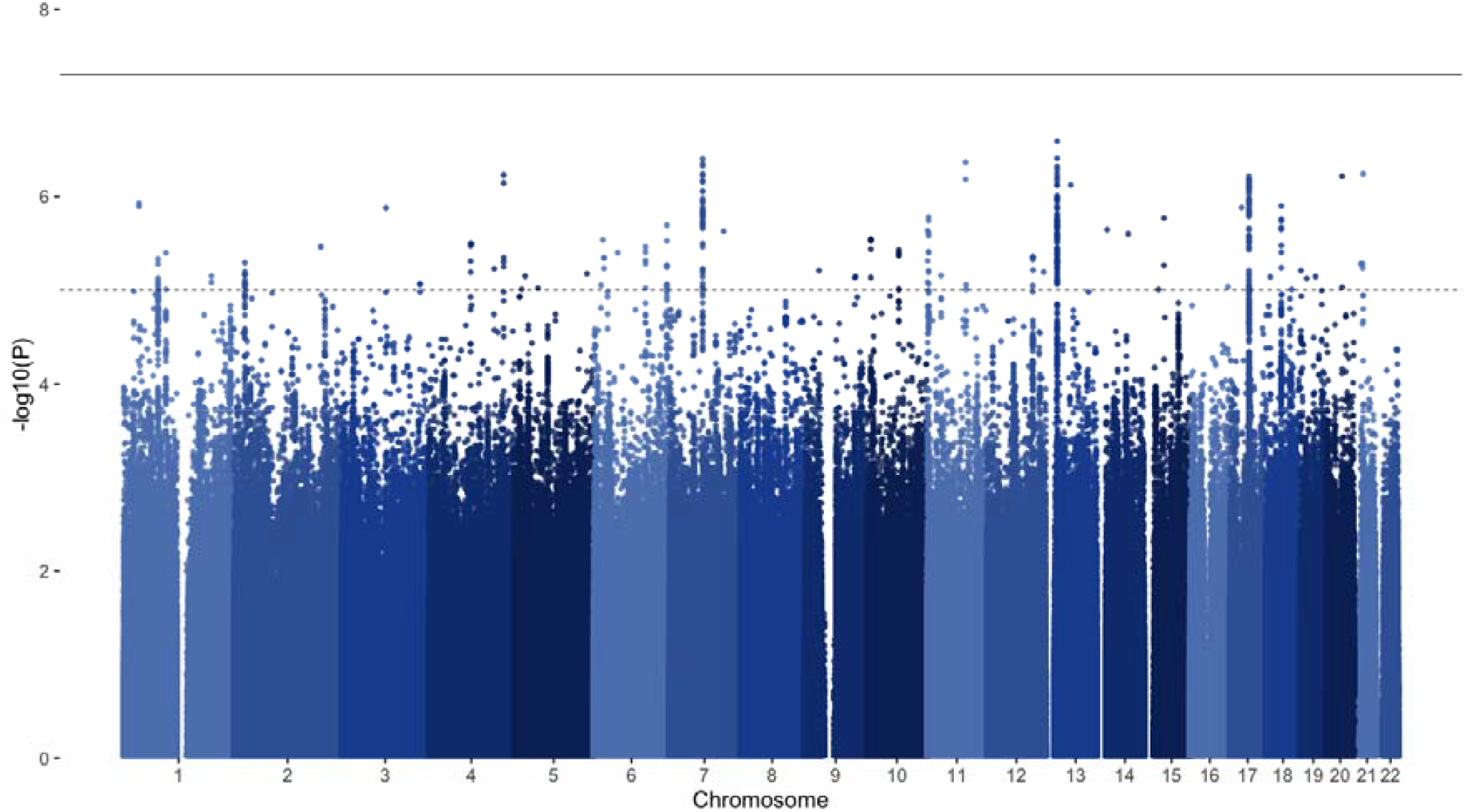
Manhattan plot of overall meta-analysis for childhood and adolescent internalising symptoms (INT_overall_). The solid line represents the significance threshold (*p* < 5×10^−08^) and the dotted line represents the suggestive threshold (*p* < 1×10^−05^).

### Stratified SNP heritabilities and within-trait genetic correlations

Estimates of SNP heritability from stratified meta-analyses are shown in Figure 2 and Supplementary Table 8. In rater-specific meta-analyses, self-reported internalising symptoms showed the highest heritability (5.63%; 95% CI 3.08–8.18%), followed by teacher, maternal, and parental report, which were all significant. Although father-reported internalising symptoms had the highest SNP heritability in rater-specific analyses (8.98%), the wide confidence intervals overlapped zero (−0.06–18.02%). In age-specific meta-analyses, SNP *h*^*2*^ for internalising symptoms in adolescence was highest (1.97%, 95% CI 0.30– 3.64%), whereas estimates for early childhood, mid-childhood, and late childhood were similar, but not robust to significance testing. In rater-by-age meta-analyses, self-reported internalising symptoms during adolescence showed the highest SNP *h*^*2*^ (3.20%, 95% CI 0.34–6.06%). Instrument-specific meta-analyses showed that variance in internalising symptoms explained by ASEBA and SDQ scales were comparable, ∼3%. The estimate for Rutter was smaller (.3%), but the difference was not substantial, based on the overlapping confidence intervals.

**Figure 2.**
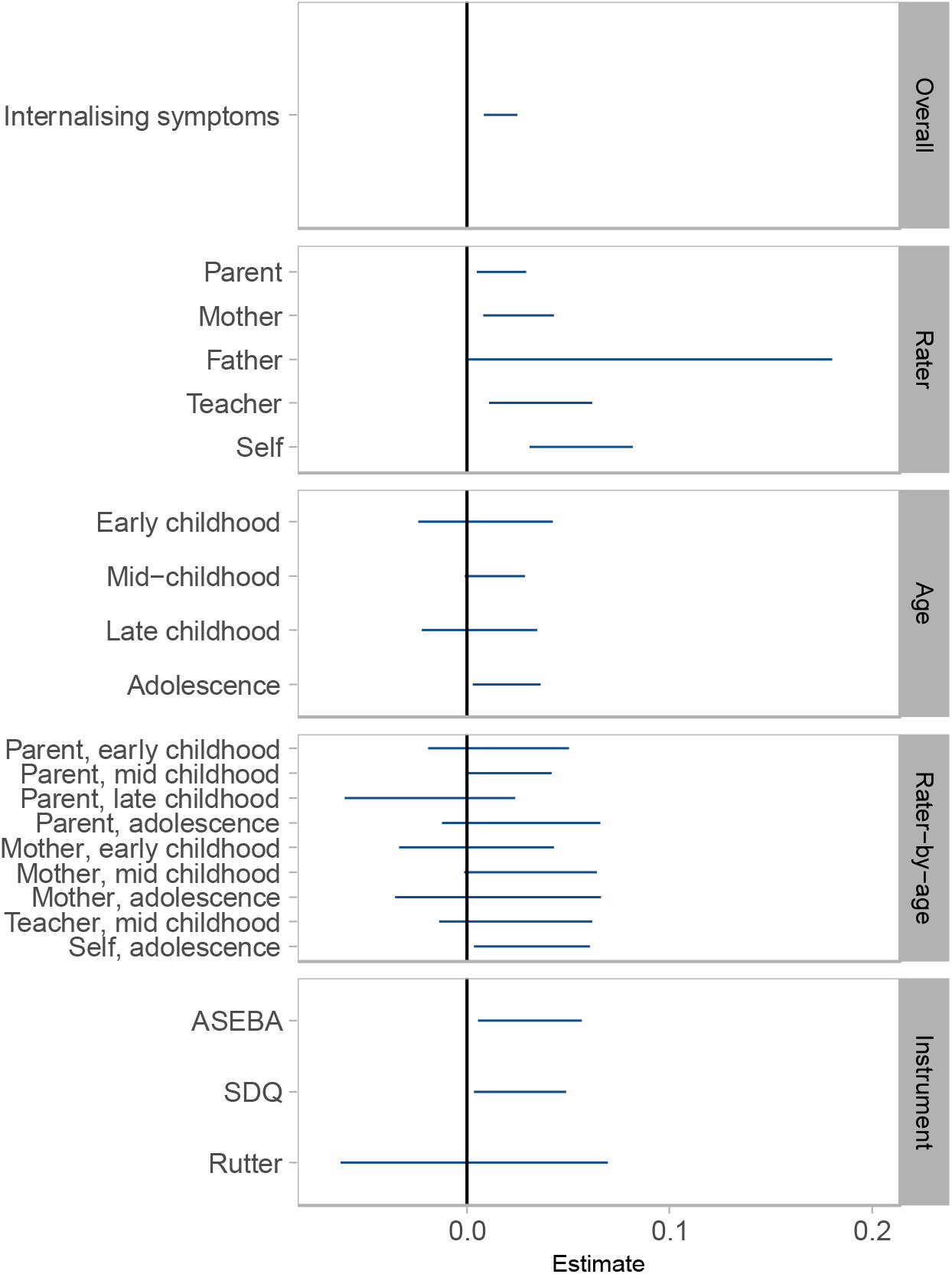
SNP heritabilities based on N-weighted meta-analyses of internalising symptoms. Error bars represent 95% confidence intervals.

INT_overall_ and self-reported internalising symptoms were highly genetically correlated (*r*_*g*_= 0.84, SE= 0.12, *p*=2.08×10^−12^). The other stratified meta-analyses were insufficiently powered to estimate genetic correlations (heritability z-score < 4).

### Gene-based analysis, tissue expression and gene-set analyses

The genome-wide gene-based analysis did not reveal any genes significantly associated with internalising symptoms, but the top 10 genes are reported in Supplementary Table 10. MAGMA tissue expression analyses of 30 general and 53 specific tissue types did not show any statistically significant associations with internalising symptoms (Supplementary Table 11). The gene-set analysis did not show any significant associations (Supplementary Table 12).

### Genetic correlations with external traits

Genetic correlations between INT_overall_ and INT_self_ (for which the z-score of the *h*^*2*^ was ≥ 4(37)), and a set of preselected external traits are shown in Figure 3 and Supplementary Table 13. INT_overall_ held strong positive genetic correlations (*r*_*g*_ *>* 0.7) with Major Depressive Disorder, anxiety, and neuroticism, and a strong negative correlation (*r*_*g*_ < -0.7) with the well-being spectrum. High correlations (|*r*_*g*_| > 0.5) with other adult and childhood psychiatric and psychological traits, including attention-deficit hyperactivity disorder (ADHD), autism spectrum disorder (ASD), depressive symptoms, loneliness, and overall and maternal-reported aggression were found. Moderate genetic correlations (|*r*_*g*_| > 0.3) with insomnia, age at first birth, cigarettes per day, educational attainment, and intelligence were also observed. INT_self_ showed a similar pattern, but generally weaker genetic associations with external traits, with some exceptions. ASD, overall and maternal-reported aggression, age at first birth, and intelligence were correlated with INT_overall_, but showed weaker correlations with INT_self_, whereas self-reported aggression, smoking initiation and body-mass index (BMI) were correlated with INT_self_, but showed weaker or no correlation with INT_overall_.

**Figure 3.**
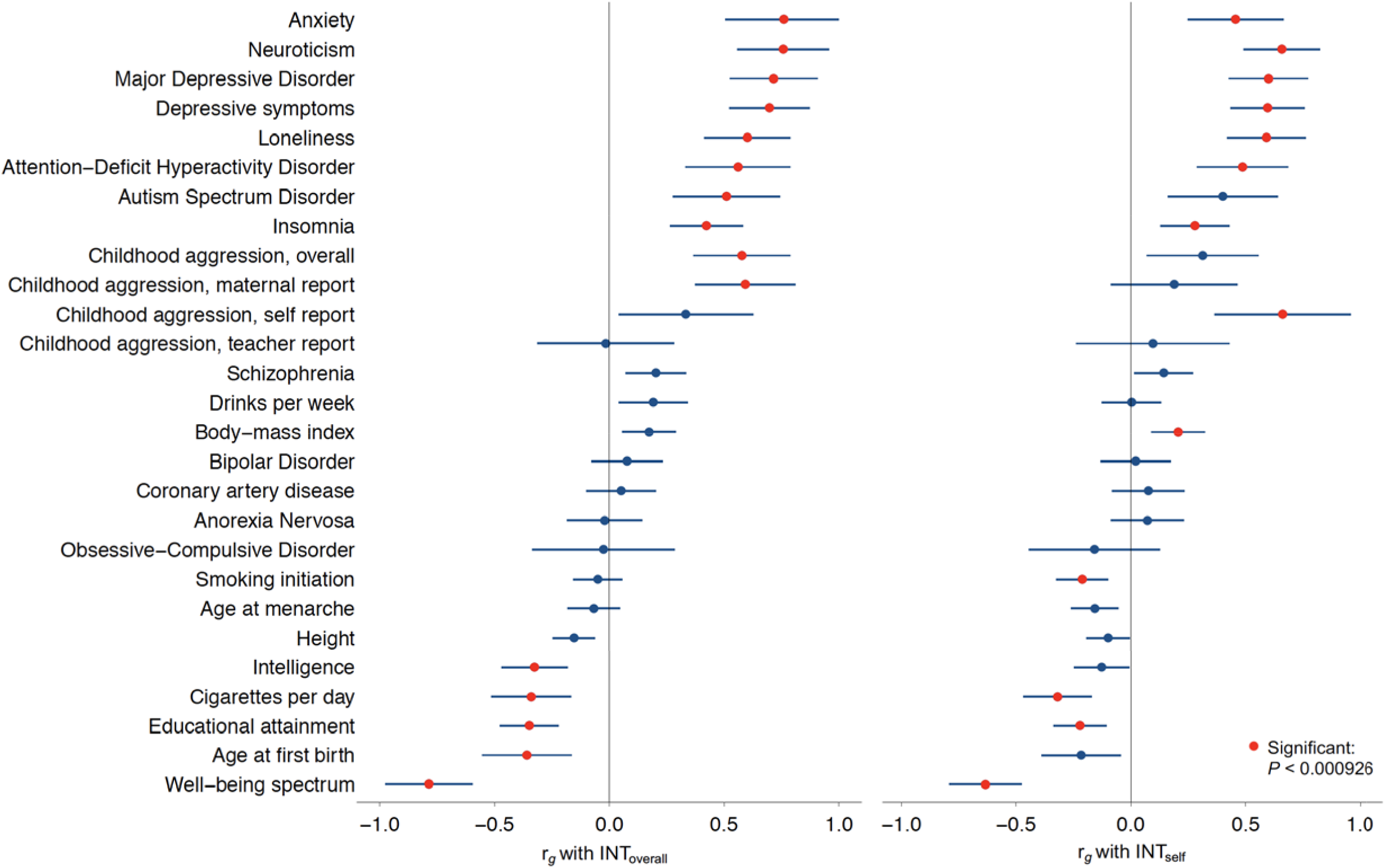
Genetic correlations with external phenotypes. The left panel shows genetic correlations with the meta-analysis for overall int ernalising symptoms in childhood and adolescence (INT_overall_) and the right panel shows genetic correlations with self-reported internalising symptoms (INT_self_). Error bars represent 95% confidence intervals. Correlation points in red are statistically significant after correction for multiple testing.

### Polygenic score prediction

Prediction of internalising symptoms in childhood and adolescence by polygenic scores based on INT_overall_ are shown in Supplementary Table 14. After correction for multiple testing, polygenic scores for INT_overall_ (N_eff_=132,260) were significantly associated with maternal-reported internalising problems in 7-year-olds, and explained up to 0.38% of the phenotypic variance. Polygenic scores for INT_overall_ were not associated with self-reported internalising problems in adolescence.

## Discussion

This genome-wide association meta-analysis of childhood and adolescent internalising symptoms included data from 64,641 individuals aged between 3 and 18. No genome-wide significant loci were detected at single SNP level. Genetic correlations with external traits suggested that childhood and adolescent internalising symptoms share substantial genetic vulnerabilities with adult internalising disorders and other childhood psychiatric traits, which could partially explain both the persistence of internalising symptoms over time and the high comorbidity amongst childhood psychiatric traits. The overall pattern of results pointed to rater-based heterogeneous effects (as discussed further on), which suggests that in addition to further increases of sample size, approaches that reduce heterogeneity will be essential in future GWAS investigations. As no biological pathways were identified, this study does not provide leads for new targets for treatment.

The most striking findings of this study are the direction and strength of genetic correlations with external traits, which point to an overlapping genetic architecture between internalising symptoms and other traits. This may initially be surprising given the low SNP heritability observed here, but while SNP heritability estimates the overall variance in a trait explained by genome-wide SNPs, a genetic correlation reflects the extent to which the same set of genetic factors are involved in two traits. As such, even traits with low SNP heritability can have high genetic correlations if the underlying set of genetic factors influencing the traits are overlapping. Strong genetic correlations (|*r*_*g*_| > 0.7) with adult depression, anxiety, neuroticism, and the wellbeing spectrum were of note, and suggest a substantial shared genetic etiology between childhood internalising symptoms and adult internalising disorders and related traits, that has also been observed in previous studies (41-43). Viewed in combination with the overlapping estimates of SNP heritability from early-childhood to adolescence in this study, these findings point to a stable set of genetic factors that partially explain the persistence of symptoms over time. Comparisons with other psychiatric disorders showed high genetic correlations (|*r*_*g*_| > 0.5) with childhood-onset disorders ADHD and ASD, but no robust associations with bipolar disorder, obsessive-compulsive disorder, or anorexia nervosa. A small genetic correlation with schizophrenia was observed (*r*_*g*_=0.2, p=.0025), which, albeit not significant due to the strict correction for multiple testing applied here, is in line with previous studies showing successful prediction of internalising symptoms in childhood using polygenic scores for schizophrenia (43-46). The overall pattern of genetic correlations with other psychiatric traits is comparable to adult cross-disorder genetic correlations, where depression shows stronger associations with ADHD and ASD than with schizophrenia or bipolar disorder (47). It appears that like adult depression, the broader (and perhaps also milder) symptomatology captured by dimensional measures of childhood internalising symptoms shares fewer genetic similarities with severe and less common disorders such as schizophrenia, bipolar disorder, OCD, and anorexia, but is more closely tied to childhood-onset disorders ADHD and ASD. This also resembles findings from the recent GWAS of total child psychiatric problems, which similarly found no robust genetic correlations with less common disorders (48). Correlations with other traits, including insomnia, loneliness, intelligence, educational attainment, cigarettes per day, and age at first birth were observed, as also seen in GWASs of adult depression and anxiety (23, 24), but unlike adult depression, no robust associations with coronary artery disease, BMI, smoking initiation, or age at menarche were found. On the other hand, both BMI and smoking initiation held robust associations with INT_self_, for which ratings were only available during adolescence. This could indicate that genetic factors during adolescence are particularly important in these associations. Age-specific genetic effects may also explain why coronary artery disease was not associated with INT_overall_, in contrast to the small but robust genetic correlation that coronary artery disease shares with both adult depression and anxiety (23, 24). This may point to genetic innovation (the involvement of novel genetic variants) in adulthood which could explain the genetic commonalities between adult internalising disorders and coronary artery disease. Alternatively, the lack of genetic correlation between INT_overall_ and coronary artery disease, as well as age of menarche (which also genetically correlates with adult depression), could be due to a lack of power. This is also shown by the wide confidence intervals for some genetic correlations (Figure 3), which can be a consequence of low SNP heritability.

Focusing on childhood traits, as well as sharing high genetic correlations with childhood-onset disorders ADHD and ASD, internalising symptoms were also highly correlated with childhood aggression. The high correlations observed across childhood traits indicate the presence of specific genetic effects that are common between childhood disorders within the neurodevelopmental spectrum. These shared genetic effects could partially explain the high comorbidity between psychiatric traits in childhood (49-51). In further examining the association between childhood internalising symptoms and aggression, INT_overall_ shared high genetic correlations with overall and maternal-reported aggression, but not with teacher or self-report. On the other hand, self-reported aggression and self-reported internalising symptoms were highly correlated, whereas INT_self_ did not share robust associations with overall, teacher, or maternal reported aggression. These patterns of rater-stratified genetic correlations suggest that observed genetic effects on childhood phenotypes can vary substantially due to differences in the phenotype captured by different raters, with the same set of raters showing the highest correlation between traits.

The difficulty in identifying causal loci for early-life internalising symptoms is not novel and resembles the trajectory of GWAS investigations of adult internalising disorders. GWAS studies of adult depression also made slow progress due to limited sample sizes and heterogeneity (52-54). As depression has several potential sources of heterogeneity, including a diverse presentation of symptoms, large case-control sample sizes were required to achieve success in identifying specific genomic loci (23, 25). GWAS studies of anxiety similarly saw increased success as sample sizes grew (24, 55). However, here we found that a substantial increase in sample size in comparison to previous studies (15, 16) did not yield the expected increase in power. In addition to heterogeneity due to a broad symptomatology, our findings indicate that GWAS investigations of childhood internalising symptoms are further disadvantaged by rater-based heterogeneous effects. Unlike adult studies where measurements are typically self or clinician reports, childhood studies, particularly those focusing on early childhood, rely heavily on parent and teacher report, which act as an additional source of heterogeneity. Rater-based differences in genetic correlations with external traits have been discussed above. The current study also observed varying estimates of SNP-heritability in rater-stratified analyses (Figure 2). Although these estimates did not appear significantly different (likely due to sample size limitations), the partial genetic correlation between INT_overall_ and INT_self_ points to incomplete overlap in relevant SNPs, indicating the presence of rater-specific genetic effects. Additionally, polygenic scores based on INT_overall_ did not predict self-reported internalising symptoms in the NTR cohort, which could also indicate heterogeneity between the target and discovery traits (56). Rater-specific genetic effects and rater disagreement on internalising symptoms are noted in previous research (57-60) and rater-based heterogeneity is also reported in the GWAS of childhood aggression (35). Heterogeneity in this study is further reflected in the low LDSC-estimate of SNP heritability in the INT_overall_ meta-analysis. This can partly be explained by the methods; estimates of SNP heritability from summary statistics are typically lower than estimates from raw genotypic data and potential over-correction of biases in LDSC could have led to more conservative estimates. However, heritability estimates rose when analyses were stratified, indicating that heterogeneity also diluted overall SNP effects in the meta-analysis.

Heterogeneous effects underlying childhood internalising symptoms can be accounted for in multivariate GWAS approaches, but our study shows that current childhood samples seem unable to meet the power requirements of these types of analyses. Another way of reducing heterogeneity and helping signal detection is to focus on diagnoses. The case-control approach has proven to be more successful than dimensional measures in adult studies of depression and anxiety (23, 24) and overcomes the limitations of treating symptom scales as continuous traits. However, diagnostic data are currently not available for childhood phenotypes in large enough samples. Instead, we expect that reducing heterogeneity at phenotypic level will be key in paving the way to success in future GWAS investigations in childhood samples. This could be tackled by examining symptom level phenotypes or separating childhood anxiety and depression into two distinct phenotypes. Another promising approach would be to eliminate heterogenous effects entirely through factor analysis. Factor analysis can be used to derive a stable phenotype which captures the core behaviour that multiple measurements (e.g. from different informants or at different time points) have in common. This eliminates variability from rater, age, or situational effects. Evidence from both twin and molecular research shows that focusing on the common part of multiple assessments results in a more reliable phenotype which shows higher heritability than that captured by individual measurements separately (41, 59, 61, 62). This way of managing rater bias has broader applicability in genetic studies within child psychiatry, but is dependent on the availability of multiple informants on behaviour at one time point.

To conclude, in this large GWAS of childhood and adolescent internalising symptoms in population-based cohorts of European ancestry, no individual loci with strong associations with the outcome were detected. However, strong genetic correlations with adult internalising traits and childhood psychiatric traits indicate that there is signal buried in the noise. Future GWAS success is likely to lie in reducing heterogeneity in childhood samples by focusing on a more stable phenotype of internalising symptoms. Finally, an important goal for future GWASs is the funding and inclusion of multi-ancestry cohorts to allow better representation of diverse populations and ensure broader applicability of findings.

## Data Availability

Summary statistics are available upon request.

https://osf.io/edas6

https://www.eagle-consortium.org/

https://www.ebi.ac.uk/gwas/

https://www.omim.org

## Notes

### Competing Interest Statement

H Larsson has served as a speaker for Evolan Pharmaand Shire/Takeda and has received research grants from Shire/Takeda, all outside the submitted work. All other authors declare no conflicts of interest.

### Clinical Protocols

https://osf.io/edas6

### Funding Statement

We extend a warm thank you to all participants, their parents, and teachers for taking part in this study. The study was supported by the Childhood and Adolescence Psychopathology: unravelling the complex etiology by a large Interdisciplinary Collaboration in Europe project (CAPICE). CAPICE received funding from the European Union Horizon 2020 research and innovation programme, Marie Sklodowska Curie Actions (MSCA ITN 2016) Innovative Training Networks, under grant agreement number 721567. Author and cohort-specific acknowledgements and funding information are described in the Supplementary Text.

### Author Declarations

Ethical approval was provided by local committees at cohort level (details are provided in the Supplementary Note). Summary statistics, with no individual data, were meta-analysed across all cohorts.

### Summary of Updates

Gene-based analysis updated, author details updated, supplementary files updated

