## Supplementary Note for "Genome-wide association meta-analysis of childhood and adolescent internalising symptoms"

### **Acknowledgments and Funding Information**

We extend a warm thank you to all participants, their parents, and teachers for taking part in this study. The project was supported by the “Childhood and Adolescence Psychopathology: unravelling the complex etiology by a large Interdisciplinary Collaboration in Europe” project (CAPICE). CAPICE received funding from the European Union’s Horizon 2020 research and innovation programme, Marie Sklodowska Curie Actions – MSCA-ITN-2016 – Innovative Training Networks, under grant agreement number 721567.

#### Author acknowledgements

AAS was supported by a NARSAD Young Investigator Award. AH was supported by the South-Eastern Norway Regional Health Authority (2018059) at Nic Waals Institute, Lovisenberg Diaconal Hospital. ARH is supported by the Children’s Hospital Foundation and University of Queensland strategic funding. AJOW is supported by an Investigator Grant from the National Health and Medical Research Council. CE and SW are supported by AG046938. CH is supported by DA042755, DA035804 and DA032555. DD is supported by a mid-career award NIH K02 AA018755, NIH R01 AA015416 (Finnish Twin Study) , P50 AA022537 (Alcohol Research Center), and R25 AA027402 (VCU GREAT), and U10 AA008401 (COGA) from the National Institute on Alcohol Abuse and Alcoholism (NIAAA). EY is supported by the Research Council of Norway grant numbers 262177 and 288083. ESJ was supported by an Academy Ter Meulen grant from the Royal Netherlands Academy of Arts and Sciences. FAH was supported by the Aggression in Children: Unraveling gene-environment interplay to inform Treatment and InterventiON strategies project (ACTION). ACTION received funding from the European Union Seventh Framework Program (FP7/2007-2013) under grant agreement no602768. JK is supported by the Academy of Finland (grants 308248, 312073). JKH, KK, AS, and MS are supported by DA011015 and DA035804. KPH is a Faculty Research Associate of the Population Research Center at the University of Texas at Austin (NICHD P2CHD042849). LME is supported by MH100141, DA044283 and AG046938. MB is supported by an ERC Consolidator Grant (WELL-BEING 771057). MGN is supported by ZonMW grants 849200011 and 531003014 937 from The Netherlands Organisation for Health Research and Development, a Jacobs Foundation Research Fellowship, and a VENI grant awarded by NWO (VI.Veni.191G.030). MRM and HMS are members of the MRC Integrative Epidemiology Unit at the University of Bristol (MC_UU_00011/7). NV-T is funded by a post-doctoral grant, Juan de la Cierva Programme (FJC2018-038085-I), Ministerio de Ciencia, Innovación y Universidades – Spanish State Research Agency. Her research has received additional support of “la Caixa” Foundation (LCF/PR/GN17/10300004) and the Health Department of the Catalan Government (Health Research and Innovation Strategic Plan (PERIS) 2016-2020 grant# SLT002/16/00201). She also acknowledges the support of the Spanish Ministry of Science, Innovation and Universities to the EMBL partnership, the Centro de Excelencia Severo Ochoa and the CERCA Programme/Generalitat de Catalunya. PL is funded by the Swedish Research Council for Health, Working Life and Welfare (project 2012-1678) and the Swedish Research Council (2016-1989). PRM is supported by the European Research Council (AdG #293574), the Bergen Research Foundation (“Utilizing the Mother and Child Cohort and the Medical Birth Registry for Better Health”), Stiftelsen Kristian Gerhard Jebsen (Translational Medical Center), the University of Bergen, the Research Council of Norway (FRIPRO grant #240413), the Western Norway Regional Health Authority (Strategic Fund “Personalized Medicine for Children and Adults”), the Novo Nordisk Foundation (grant #54741), and the Norwegian Diabetes Association. PM is supported by the Research Council of Norway through its Centers of Excellence scheme, project no 262700. RB was supported by NIH grants (MH100141, MH016880). REP is supported by the National Institutes of Health K01MH113848 and The Brain & Behavior Research Foundation Young Investigator Grant 28632. RP is supported by a Medical Research Council Professorship award (G19/2). KR is supported by a Sir Henry Wellcome Postdoctoral Fellowship. RPC is supported by DA011015, AG046938 and DA035804. RR was supported by a Research Scientist Award from NIH (AA-000145). SA is funded by a Juan de la Cierva – Incorporación Postdoctoral Contract from Ministerio de Economía, Industria y Competitividad (IJCI-2017-34068). SAB is supported by DA035804. SL is also supported by the Swedish Research Council (2017-02552). TLW is supported by DA03580 and DA021905. TR-K is supported by the Research Council of Norway grant #274611. WC is supported by MH117559 and HD093651.

## 1958BC

This work made use of data and samples generated by the 1958 Birth Cohort (NCDS), which is managed by the Centre for Longitudinal Studies at the UCL Institute of Education. The authors are deeply grateful to the 1958 birth cohort participants for their longstanding commitment and support, and to all staff for cohort coordination and data collection. Statistical analyses were funded by National Health and Medical Research Council, Australia (GNT1157281). The management of the 1958 Birth Cohort is funded by the Economic and Social Research Council (grant number ES/M001660/1). Access to these resources was enabled via the 58READIE Project funded by Wellcome Trust and Medical Research Council (grant numbers WT095219MA and G1001799). DNA collection was funded by MRC grant G0000934 and cell-line creation by Wellcome Trust grant 068545/Z/02. This study makes use of data generated by the Wellcome Trust Case-Control Consortium. A full list of investigators who contributed to generation of the data is available from the Wellcome Trust Case-Control Consortium website. Funding for the project was provided by the Wellcome Trust under the award 076113. This research used resources provided by the Type 1 Diabetes Genetics Consortium, a collaborative clinical study sponsored by the National Institute of Diabetes and Digestive and Kidney Diseases (NIDDK), National Institute of Allergy and Infectious Diseases, National Human Genome Research Institute, National Institute of Child Health and Human Development, and Juvenile Diabetes Research Foundation International (JDRF) and supported by U01 DK062418.

#### ABCD

Data used in the preparation of this article were obtained from the Adolescent Brain Cognitive Development (ABCD) Study (<https://abcdstudy.org>), held in the NIMH Data Archive (NDA). The ABCD Study is supported by the National Institutes of Health (NIH) and additional federal partners under award numbers U01DA041022, U01DA041025, U01DA041028, U01DA041048, U01DA041089, U01DA041093, U01DA041106, U01DA041117, U01DA041120, U01DA041134, U01DA041148, U01DA041156, U01DA041174, U24DA041123, and U24DA041147. A full list of federal partners is available at <https://abcdstudy.org/federal-partners.html>. A listing of participating sites and a complete listing of the study investigators can be found at <https://abcdstudy.org/principal-investigators.html>. This manuscript reflects the views of the authors and may not reflect the opinions or views of the NIH or ABCD consortium investigators. ABCD consortium investigators designed and implemented the study and/or provided data but did not necessarily participate in analysis or writing of this report. The ABCD data repository grows and changes over time. The ABCD data used in this report came from Data Release 2.0.1 (doi: [10.15154/1504041](http://dx.doi.org/10.15154/1504041)).

#### Add Health

We gratefully acknowledge research support from the National Institute of Child Health and Human Development to Harris through grant 3 P01 HD31921 as part of the Add Health program project. This research uses data from Add Health, a program project designed by J. Richard Udry, Peter S. Bearman, and Kathleen Mullan Harris, and funded by a grant P01-HD31921 from the National Institute of Child Health and Human Development, with cooperative funding from 23 other agencies. Add Health GWAS data were funded by NICHD Grants R01 HD073342 (Harris) and R01 HD060726 (Harris, Boardman, and McQueen). Information on how to obtain Add Health data files is available on the Add Health website (<http://www.cpc.unc.edu/addhealth>). We thank the Add Health participants for making this research possible.

#### ALSPAC

We are extremely grateful to all the families who took part in this study, the midwives for their help in recruiting them, and the whole ALSPAC team, which includes interviewers, computer and laboratory technicians, clerical workers, research scientists, volunteers, managers, receptionists and nurses. The UK Medical Research Council and Wellcome (Grant ref: 102215/2/13/2) and the University of Bristol provide core support for ALSPAC. This publication is the work of the authors who will serve as guarantors for the contents of this paper. A comprehensive list of grants funding is available on the ALSPAC website (http://www.bristol.ac.uk/alspac/external/documents/grant-acknowledgements.pdf). GWAS data was generated by Sample Logistics and Genotyping Facilities at Wellcome Sanger Institute and LabCorp (Laboratory Corporation of America) using support from 23andMe. This study was supported by the National Institute for Health Research (NIHR) Biomedical Research Centre at the University Hospitals Bristol National Health Service Foundation Trust and the University of Bristol. The views expressed in this publication are those of the author(s) and not necessarily those of the National Health Service, the National Institute for Health Research or the Department of Health.

#### BREATHE

This cohort was funded by the European Research Council under the ERC Grant Agreement number 268479 – the BREATHE project. ISGlobal acknowledge support from the Spanish Ministry of Science and Innovation through the “Centro de Excelencia Severo Ochoa 2019-2023” Program (CEX2018-000806-S), and support from the Generalitat de Catalunya through the CERCA Program. We acknowledge all the children and their families participating into the BREATHE project for their altruism.

#### CATSS

The Child and Adolescent Twin Study in Sweden study was supported by the Swedish Council for Working Life, funds under the ALF agreement, the Söderström Königska Foundation and the Swedish Research Council (Medicine, Humanities and Social Sciences, and SIMSAM). We acknowledge The Swedish Twin Registry for access to data which is managed by Karolinska Institutet and receives funding through the Swedish Research Council under the grant no 2017-00641.

#### FinnTwin12

We wish to thank all twins and their families for their participation. Data collection has been supported by the National Institute of Alcohol Abuse and Alcoholism (grants AA-12502, AA-00145, and AA-09203 to Richard J Rose) and the Academy of Finland (grants 100499, 205585, 118555, 141054 and 264146 to Jaakko Kaprio).

#### GSMS

This research was supported by the National Institute on Drug Abuse (U01DA024413, R01DA11301), the National Institute of Mental Health (R01MH063970, R01MH063671, R01MH048085, K01MH093731 and K23MH080230), NARSAD, and the William T. Grant Foundation. We are grateful to all the GSMS and CCC study participants who contributed to this work.

#### Gen-R

This work was supported by the Dutch Ministry of Education, Culture and Science (Gravity Grant No. 024.001.003, Consortium on Individual Development), and the Netherlands Organisation for Scientific Research (NWO‐grant 016.VICI.170.200) to HT. The first phase of the Generation R Study is made possible by financial support from the Erasmus Medical Centre, Rotterdam; the Erasmus University Rotterdam; and the Netherlands Organisation for Health Research and Development (ZonMw). The authors gratefully acknowledge the contribution of all children and parents, general practitioners, hospitals, midwives and pharmacies involved in the Generation R Study. The Generation R Study is conducted by the Erasmus Medical Centre (Rotterdam) in close collaboration with the School of Law and Faculty of Social Sciences of the Erasmus University Rotterdam; the Municipal Health Service Rotterdam area, Rotterdam; the Rotterdam Homecare Foundation, Rotterdam; and the Stichting Trombosedienst & Artsenlaboratorium Rijnmond, Rotterdam.

GINIplus
The GINIplus study was mainly supported for the first 3 years of the Federal Ministry for Education, Science, Research and Technology (interventional arm) and Helmholtz Zentrum Munich (former GSF) (observational arm). The 4 year, 6 year, 10 year and 15 year follow-up examinations of the GINIplus study were covered from the respective budgets of the 5 study centres (Helmholtz Zentrum Munich (former GSF), Research Institute at Marien-Hospital Wesel, LMU Munich, TU Munich and from 6 years onwards also from IUF - Leibniz Research-Institute for Environmental Medicine at the University of Düsseldorf) and a grant from the Federal Ministry for Environment (IUF Düsseldorf, FKZ 20462296). Further, the 15 year follow-up examination of the GINIplus study was supported by the Commission of the European Communities, the 7th Framework Program: MeDALL project, and as well by the companies Mead Johnson and Nestlé.

LISA
The LISA study was mainly supported by grants from the Federal Ministry for Education, Science, Research and Technology and in addition from Helmholtz Zentrum Munich (former GSF), Helmholtz Centre for Environmental Research - UFZ, Leipzig, Research Institute at Marien-Hospital Wesel, Pediatric Practice, Bad Honnef for the first 2 years. The 4 year, 6 year, 10 year and 15 year follow-up examinations of the LISA study were covered from the respective budgets of the involved partners (Helmholtz Zentrum Munich (former GSF), Helmholtz Centre for Environmental Research - UFZ, Leipzig, Research Institute at Marien-Hospital Wesel, Pediatric Practice, Bad Honnef, IUF – Leibniz-Research Institute for Environmental Medicine at the University of Düsseldorf) and in addition by a grant from the Federal Ministry for Environment (IUF Düsseldorf, FKZ 20462296). Further, the 15-year follow-up examination of the LISA study was supported by the Commission of the European Communities; MeDALL project.

#### IBG

Funding for genotyping, analytic support, and data curation supported by NIH R01 AG046938 and P60 DA011015.

#### MOBA

The Norwegian Mother, Father and Child Cohort Study is supported by the Norwegian Ministry of Health and Care Services and the Ministry of Education and Research. We are grateful to all the participating families in Norway who take part in this on-going cohort study. We thank the Norwegian Institute of Public Health (NIPH) for generating high-quality genomic data. This research is part of the HARVEST collaboration, supported by the Research Council of Norway (#229624). We also thank the NORMENT Centre for providing genotype data, funded by the Research Council of Norway (#223273), South East Norway Health Authority and KG Jebsen Stiftelsen. We further thank the Center for Diabetes Research, the University of Bergen for providing genotype data funded by the ERC AdG project SELECTionPREDISPOSED, Stiftelsen Kristian Gerhard Jebsen, Trond Mohn Foundation, the Research Council of Norway, the Novo Nordisk Foundation, the University of Bergen, and the Western Norway health Authorities (Helse Vest).

#### MSUTR

This research was supported by the National Institute of Mental Health (NIMH) under Award Number R01-MH081813 and the Eunice Kennedy Shriver National Institute for Child Health and Human Development (NICHD) under Award Number R01-HD066040. This research was supported by the National Institute on Drug Abuse under Award Number R01DA043501 and the National Library of Medicine under Award Number R01LM012848.

#### MUSP

This study was funded by grants received from the National Health and Medical Research Council (NHMRC) and Australian Research Council (ARC). Thanks also to Shelby Marrington (Project Manager) and Greg Shuttlewood (Data Manager) who have supervised the day-to-day management of the study. We also extend our thanks to the mothers and children who have continued to participate in the study.

#### NFBC

We thank all cohort members and researchers who have participated in the study. We also wish to acknowledge the work of the NFBC project center. NFBC1986 has received funding from: EU QLG1-CT-2000-01643 (EUROBLCS) Grant no. E51560, NorFA Grant no. 731, 20056, 30167, USA / NIHH 2000 G DF682 Grant no. 50945, the EU H2020-MSCA-ITN-2016 CAPICE Action Grant no. 721567. Academy of Finland EGEA project (285547), EU H2020 LifeCycle Action (grant agreement No 733206), and DynaHEALTH action (grant agreements No. 633595). The DNA extractions, sample quality controls, biobank upkeep and aliquoting were performed in the National Public Health Institute, Biomedicum Helsinki, Finland and supported financially by the Academy of Finland and Biocentrum Helsinki.

#### NTR

Funding was obtained from multiple grants from the Netherlands Organization for Scientific Research (NWO) and The Netherlands Organisation for Health Research and Development (ZonMW): Genetic influences on stability and change in psychopathology from childhood to young adulthood (ZonMw 912-10-020); Twin family database for behavior genomics studies (NWO 480-04-004); Genetic and Family Influences on Adolescent. Psychopathology and Wellness (NWO 463-06-001); A Twin-Sibling Study of Adolescent Wellness (451-04-034). Twin research focusing on behavior (NWO 400-05-717); Longitudinal data collection from teachers of Dutch twins and their siblings (481-08-011), Twin-family-study of individual differences in school achievement (NWO-FES, 056-32-010), Genotype/phenotype database for behavior genetic and genetic epidemiological studies (ZonMw Middelgroot 911-09-032); “Why some children thrive” (OCW_Gravity program –NWO-024.001.003), Netherlands Twin Registry Repository: researching the interplay between genome and environment (NWO-Groot 480-15-001/674); BBMRI –NL (184.021.007 and 184.033.111): Biobanking and Biomolecular Resources Research Infrastructure; Spinozapremie (NWO- 56-464-14192) and KNAW Academy Professor Award (PAH/6635) to DIB ; the Neuroscience Campus Amsterdam (NCA) and Amsterdam Public Health (APH); the European Science Council (ERC) Genetics of Mental Illness (ERC Advanced, 230374); NIH: Rutgers University Cell and DNA Repository cooperative agreement (NIMH U24 MH068457-06); Developmental Study of Attention Problems in Young Twins (NIMH, RO1 MH58799-03); Grand Opportunity grant Developmental trajectories of psychopathology (NIMH 1RC2 MH089995), National Institute of Diabetes and Digestive and Kidney Diseases (RO1DK092127), European Union Seventh Framework Program “ACTION” - Aggression in Children: unravelling gene-environment interplay to inform Treatment and InterventiON strategies project (FP7 602768), and the Avera Institute for Human Genetics.

#### PNC

Support for the collection of the data for Philadelphia Neurodevelopment Cohort (PNC) was provided by grant RC2MH089983 awarded to Raquel Gur and RC2MH089924 awarded to Hakon Hakonarson. Subjects were recruited and genotyped through the Center for Applied Genomics (CAG) at The Children's Hospital in Philadelphia (CHOP). Phenotypic data collection occurred at the CAG/CHOP and at the Brain Behavior Laboratory, University of Pennsylvania. We thank the PNC participants for making this research possible, as well as the original study investigators for making their data publicly available.

#### The Raine Study

The Raine Study acknowledges the National Health and Medical Research Council (NHMRC) for their long term contribution to funding the study over the last 30 years. Core Management of the Raine Study has been funded by the University of Western Australia (UWA), Curtin University, the Raine Medical Research Foundation, the Telethon Kids Institute, the Women's and Infants Research Foundation, Edith Cowan University, Murdoch University, and the University of Notre Dame (Australia). This study was supported by the National Health and Medical Research Council of Australia [grant numbers 572613, 403981 and 003209] and the Canadian Institutes of Health Research [grant number MOP-82893]. We would also like to acknowledge The University of Western Australia (Division of Obstetrics and Gynaecology, King Edward Memorial Hospital and Medical School, Royal Perth Hospital), and Telethon Kids Institute for providing in-kind support for the storage and curation of biological samples. All analytic work was supported by resources provided by the Pawsey Supercomputing Centre with funding from the Australian Government and the Government of Western Australia.

#### TEDS

We gratefully acknowledge the ongoing contribution of the participants in the Twins Early Development Study (TEDS) and their families. TEDS is supported by a programme grant to RP from the UK Medical Research Council (MR/M021475/1 and previously G0901245), with additional support from the US National Institutes of Health (AG046938). The research leading to these results has also received funding from the European Research Council under the European Union’s Seventh Framework Programme (FP7/2007- 2013)/grant agreement n° 602768 and ERC grant agreement n° 295366. This study represents independent research partly funded by the NIHR BRC at South London and Maudsley NHS Foundation Trust and King’s College London. The views expressed are those of the author(s) and not necessarily those of the NHS, the NIHR or the Department of Health and Social Care. High performance computing facilities were funded with capital equipment grants from the GSTT Charity (TR130505) and Maudsley Charity (980).

#### TRAILS

TRAILS (TRacking Adolescents’ Individual Lives Survey) is a collaborative project involving various departments of the University Medical Center and University of Groningen, the University of Utrecht, the Radboud Medical Center Nijmegen, and the Parnassia Bavo group, all in the Netherlands. TRAILS has been financially supported by grants from the Netherlands Organization for Scientific Research NWO (Medical Research Council program grant GB-MW 940-38-011; ZonMW Brainpower grant 100-001-004; ZonMw Risk Behavior and Dependence grant 60-60600-97-118; ZonMw Culture and Health grant 261-98-710; Social Sciences Council medium-sized investment grants GB-MaGW 480-01-006 and GB-MaGW 480-07-001; Social Sciences Council project grants GB-MaGW 452-04-314 and GB-MaGW 452-06-004; NWO large-sized investment grant 175.010.2003.005; NWO Longitudinal Survey and Panel Funding 481-08-013 and 481-11-001; NWO Vici 016.130.002 and 453-16-007/2735; NWO Gravitation 024.001.003); the Dutch Ministry of Justice (WODC), the European Science Foundation (EuroSTRESS project FP-006), the European Research Council (ERC-2017-STG-757364 and ERC-CoG-2015-681466), Biobanking and Biomolecular Resources Research Infrastructure BBMRI-NL (CP 32), the Gratama foundation, the Jan Dekker foundation, the participating universities, and Accare Center for Child and Adolescent Psychiatry. We are grateful to all adolescents, their parents and teachers who participated in this research and to everyone who worked on this project and made it possible. Statistical analyses were carried out on the Genetic Cluster Computer (http://www.geneticcluster.org), which is financially supported by the Netherlands Scientific Organization (NWO 480-05-003) along with a supplement from the Dutch Brain Foundation.

#### VTSABD

This research was supported by the National Institute on Drug Abuse (U01DA024413, R01DA025109), the National Institute of Mental Health (R01MH045268, R01MH068521). We are grateful to all the VTSABD study participants who contributed to this work.

#### YFS

The Young Finns Study has been financially supported by the Academy of Finland: grants 322098, 286284, 134309 (Eye), 126925, 121584, 124282, 129378 (Salve), 117787 (Gendi), and 41071 (Skidi); the Social Insurance Institution of Finland; Competitive State Research Financing of the Expert Responsibility area of Kuopio, Tampere and Turku University Hospitals (grant X51001); Juho Vainio Foundation; Paavo Nurmi Foundation; Finnish Foundation for Cardiovascular Research ; Finnish Cultural Foundation; The Sigrid Juselius Foundation; Tampere Tuberculosis Foundation; Emil Aaltonen Foundation; Yrjö Jahnsson Foundation; Signe and Ane Gyllenberg Foundation; Diabetes Research Foundation of Finnish Diabetes Association; EU Horizon 2020 (grant 755320 for TAXINOMISIS and grant 848146 for TO AITION); European Research Council (grant 742927 for MULTIEPIGEN project); and Tampere University Hospital Supporting Foundation. We thank the teams that collected data at all measurement time points; the persons who participated as both children and adults in these longitudinal studies; and biostatisticians Irina Lisinen, Johanna Ikonen, Noora Kartiosuo, Ville Aalto, and Jarno Kankaanranta for data management and statistical advice.

### **Cohort Descriptions**

## 1958BC

The 1958 British Birth Cohort (1958BC) covered all births during one week in March 1958 in England, Scotland, and Wales (n=17,638) (Power & Elliott, 2006). Ethical approval for the 45 years survey was obtained from the South East Multicenter Research Ethics Committee (ref. 01/1/44) and the Joint UCL/UCLH Committees on the Ethnics of Human Research (Committee A) (ref. 08/H0714/40). Childhood follow-ups were conducted at age 7 and 11 years, when parents also rated children’s emotional difficulties using short forms of Rutter, Tizard & Whitmore’s Health and Behaviour Checklists (Rutter, Tizard, & Whitmore, 1970). Genome-wide data for the 1958BC has been obtained through two sub-studies using the 1958BC as a source for population controls: 3,000 samples were randomly selected as part of the Wellcome Trust Case Control Consortium (Sawcer et al., 2011) and 2,592 distinct samples were selected as part of the Type 1 Diabetes Genetics Consortium (Barrett et al., 2009).

#### ABCD

The Adolescent Brain Cognitive Development Study (ABCD) is an ongoing nation-wide study, comprising of 11,875 children aged 9–10 years, recruited to match national socio-economic variation. The sociodemographic factors on which the sample was recruited were age, sex, race, socioeconomic status, and urbanicity. Participants were enrolled at 22 sites, the catchment area of which encompassed over 20% of the entire US population in this age group. Multi-stage sampling was used to ensure both local randomisation and representativeness of sociodemographic variation of the ABCD sample (Garavan et al., 2018). ABCD study data is available to scientists worldwide to conduct research on the many factors that influence brain, cognitive, social, and emotional development. The data used in this study were accessed from the ABCD Study Curated Annual Release 2.0.1 and are available on request from the National Institute of Mental Health data archive. The University of California at San Diego Institutional Review Board is responsible for the ethical oversight of the ABCD study.

#### Add Health

The National Longitudinal Study of Adolescent to Adult Health (Add Health) is a large, nationally-representative longitudinal cohort study of adolescents who have been followed into adulthood for 20+ years. The Add Health cohort has extensive longitudinal data on a myriad of social, behavioural, environmental, and biological variables, and nearly 10,000 participants now have genome-wide genetic data. For more information, we refer the reader to previous publications that describe the design of the study (Harris, Halpern, Haberstick, & Smolen, 2013; Harris, Lee, & DeLeone, 2010).

#### ALSPAC

The Avon Longitudinal Study of Parents and Children (ALSPAC) is a longitudinal pregnancy cohort which aimed to recruit all pregnant women in the former county of Avon with an expected due date between April 1991 and December 1992. Detailed information has continued to be collected on mothers, partners and children from 15,454 pregnancies in the cohort, this process has been described in detail elsewhere (Boyd et al., 2013; Fraser et al., 2013; Northstone et al., 2019). Ethics approval for the study was obtained from the ALSPAC Ethics and Law Committee and the Local Research Ethics Committees. Informed consent for the use of data collected via questionnaires and clinics was obtained from participants following the recommendations of the ALSPAC Ethics and Law Committee at the time. Consent for biological samples has been collected in accordance with the Human Tissue Act (2004). Please note that the study website contains details of all the data that is available through a fully searchable data dictionary and variable search tool: <http://www.bristol.ac.uk/alspac/researchers/our-data/>.

#### BREATHE

The BREATHE project (BRain dEvelopment and Air polluTion ultrafine particles in scHool childrEn) is a population-based cohort of primary schoolchildren designed to analyse the association between air pollution and behaviour, cognitive function and brain morphology. All families of children without special needs who were enrolled in 2nd, 3rd, and 4th grades at the selected schools were invited to participate in the study. A total of 2897 children aged 7 to 11 years accepted the invitation and participated in the project. Genotype data were available for 1667 children of European ethnic origin. Further details can be found elsewhere (Alemany et al., 2016; Sunyer et al., 2015).

#### CATSS

The Child and Adolescent Twin Study in Sweden (CATSS) is an ongoing longitudinal twin study targeting all twins born in Sweden since July 1, 1992 (Anckarsäter et al., 2011; Lichtenstein, Tuvblad, Larsson, & Carlström, 2007). Parents of twins are interviewed regarding the children’s somatic and mental health and social environment in connection with their 9th or 12th birthdays and followed up over time. The response rate was 75%. Follow-ups were conducted when the twins were 15 years of age (response rate, 61%) and 18 years of age (response rate, 59%). All responding parents, legal guardians or twins provided informed consent; digitally, written or by participation, to the study, and all data were deidentified. This study received ethical approval from the Karolinska Institutet Ethical Review Board. DNA samples (from saliva) were obtained from the participants at study enrolment (Brikell et al., 2018). A total of 11 081 samples passed quality control assessment; MZ twins were then imputed, resulting in 13 576 samples and 6 981 993 imputed SNPs that passed all quality control assessments.

#### FinnTwin12

FinnTwin12 is a population-based cohort of Finnish twins born 1983–1987 that was established to follow health and behavioural habits (Kaprio, 2006, 2013; Kaprio, Pulkkinen, & Rose, 2002; Rose et al., 2019). Families of twins were identified through the Finnish Central Population Registry and initially enrolled when the twins were ages 11–12 years old (N=5600 twins; 87% response rate). Phenotype data on depressiveness and social anxiety was collected via questionnaire (Pulkkinen, Kaprio, & Rose, 1999) from multiple informants over time: from the twins themselves (including questions about their co-twin) at ages 14 and 17; from parents at age 12; and from teachers at ages 12 and 14. A total of 1479 had DNA samples (from blood or saliva) taken at age 22 for genotyping.

#### GSMS

The Great Smoky Mountains Study (GSMS) is a longitudinal, representative study of 1 420 children in 11 predominantly rural counties in Southeastern United States (Copeland, Angold, Shanahan, & Costello, 2014; Costello et al., 1996). Annual assessments on psychopathology and associated factors were completed on the 1 420 children until age 16 (6 674 observations of 1 420 individuals; 1993 to 2000) and then again at ages 19, 21, 25, and 30 (4 556 observations of 1 336 participants; 1999 to 2015) for a total of 11,230 total assessments.

#### Gen-R

The Generation R (Gen-R) study is a prospective cohort study from fetal life onwards that included pregnant women living in Rotterdam, the Netherlands, with an expected delivery date between April 2002 and January 2006 (n = 9,778). The main aim of this study is to identify early environmental and genetic factors that affect growth, health and development (Kooijman et al., 2016). The Generation R Study is multidisciplinary and both prenatal and postnatal measures have included multiple domains of growth, health and development. Rotterdam is an ethnically diverse city and this is reflected in the Generation R participants. Of the enrolled mothers, 42% was of non-Dutch ethnic background, largely made by mothers from Surinamese (9%), Turkish (7%) and Moroccan (3%) background (Kooijman et al., 2016; Medina-Gomez et al., 2015). Data has been collected in children up until the mean age of 10 years, with current on-going data collection at mean age 13 years. Study protocols were approved by the local ethics committee, and written informed consent and assent was obtained from all parents and children.

#### GINIplus

A total of 5991 mothers and their newborns were recruited into the German Infant study on the influence of Nutrition Intervention PLUS environmental and genetic influences on allergy development (GINIplus) between September 1995 and June 1998 in Munich and Wesel. Infants with at least one allergic parent and/or sibling were allocated to the interventional study arm investigating the effect of different hydrolysed formulas for allergy prevention in the first year of life (Berg et al., 2010). All children without a family history of allergic diseases and children whose parents did not give consent for the intervention were allocated to the non-interventional arm. The present study is limited to participants from the Munich study center. The phenotype was obtained from the emotional problems subscale of the Strength and Difficulties Questionnaire (SDQ), which was administered to the parents at age 10 years and participants themselves at age 15 years. DNA was collected at the age 6 and 10 years. Approval by the local Ethics Committees and written consent from participant’s families were obtained.

#### LISA

The influence of Life-style factors on the development of the Immune System and Allergies in East and West Germany PLUS the influence of traffic emissions and genetics (LISAplus) Study is a population based birth cohort study. A total of 3094 healthy, full-term neonates were recruited between 1997 and 1999 in Munich, Leipzig, Wesel and Bad Honnef (Heinrich et al., 2002). The participants were not pre-selected based on family history of allergic diseases. The present study is limited to participants from the Munich study center. The phenotype was obtained from the emotional problems subscale of the Strength and Difficulties Questionnaire (SDQ), which was administered to the parents at age 10 years and participants themselves at age 15 years. DNA was collected at the age 6 and 10 years. Approval by the local Ethics Committees and written consent from participant’s families were obtained.

#### IBG

Two cohort studies from the Institute for Behavioral Genetics (IBG) supplied information for this project: The Colorado Adoption Project (<http://ibgwww.colorado.edu/cap/>) and the Colorado Twin Registry (<https://www.colorado.edu/ibg/research/human-research-studies/colorado-twin-registry>) (Derringer et al., 2015; Rhea, Bricker, Wadsworth, & Corley, 2013; Rhea, Gross, Haberstick, & Corley, 2013).

#### MOBA

The Norwegian Mother, Father and Child Cohort Study (MoBa) is a population-based pregnancy cohort study conducted by the Norwegian Institute of Public Health (Magnus, Birke, Vejrup, Haugan, Alsaker, Daltveit, Handal, Haugen, Høiseth, & Knudsen, 2016). Participants were recruited from all over Norway from 1999-2008. The women consented to participation in 41% of the pregnancies. The cohort now includes 114,500 children, 95,200 mothers and 75,200 fathers. The current study is based on version 10 of the quality-assured data files released for research in 2018. Parent and infant DNA samples were collected at birth and stored in a biobank. Of these, 11,000 randomly-selected trios (mother, father, offspring) were genotyped as part of the HARVEST project (Magnus, Birke, Vejrup, Haugan, Alsaker, Daltveit, Handal, Haugen, Høiseth, Knudsen, et al., 2016), which is the data used in this study. The establishment of MoBa and initial data collection was based on a license from the Norwegian Data protection agency and approval from The Regional Committees for Medical and Health Research Ethics. The MoBa cohort is based on regulations based on the Norwegian Health Registry Act. The current study was approved by The Regional Committees for Medical and Health Research Ethics (REK 2013/863).

#### MSUTR

Data for this study comes from the Michigan State University Twin Registry (MSUTR), a large, population-based twin registry comprised of thousands of twins throughout Michigan, aged 3-55 (current N~31 300) (Burt & Klump, 2013). The overall focus of the MSUTR is on understanding developmental changes in genetic, environmental, and neurobiological influences on internalizing and externalizing disorders. Recruitment for the MSUTR is ongoing via the identification of birth records through the Michigan Department of Health and Human Services (MDHHS). Because birth records are confidential in Michigan, recruitment packets are mailed directly from MDHHS to eligible twin pairs. Twins indicating interest in participation via pre-stamped postcards or e-mails/calls to the MSUTR project office are then contacted by study staff to determine study eligibility and to schedule their assessments. Participants in the registry complete a family health and demographic questionnaire via mail. Families are then recruited for one or more of the intensive, in-person studies based on their answers to relevant items in the registry questionnaire. In-person assessments target a variety of biological, genetic, and environmental phenotypes, including multi-informant measures of psychiatric and behavioural phenotypes, census and neighbourhood informant reports of twin neighbourhood characteristics, buccal swab and salivary DNA samples, assays of adolescent and adult steroid hormone levels, and/or videotaped interactions of child twin families. Approval for all studies was obtained through the Michigan State University Institutional Review Board and the Michigan Department of Health and Human Services Institutional Review Board. All methods were performed in accordance to relevant guidelines and regulations.

#### MUSP

The MUSP (Mater Misericordiae Mothers' Hospital-University of Queensland Study of Pregnancy) study is of 7 223 women recruited early in pregnancy over the period 1981-1983 and the live singleton children to whom they subsequently gave birth. There were follow-ups of the mothers at 5, 14, 21 and 27 years after recruitment. Children were also followed-up in the mothers’ questionnaire and independently at 21 and 30 years of age. The CBCL and YSR were administered to children at 5, 14 and 21 years of age. The CIDI was administered to mothers at 27 years after recruitment and the children at 21 and 30 years after recruitment. The cohort comprises both mothers (to 27 years after the birth) and children (to 30 years of age). Three papers describing the recruitment methodology and sample details have been published (Keeping et al., 1989; Najman et al., 2015; Najman et al., 2005).

#### NFBC

Northern Finland Birth Cohort 1986 (NFBC1986) is a prospective longitudinal birth cohort which included pregnant women with expected date of delivery between July 1985 and June 1986 in the two Northern most provinces of Finland. In total, 9 432 children were live-born in the cohort (Järvelin, Hartikainen‐Sorri, & Rantakallio, 1993). At approximately age 16 years, the cohort members were asked to complete a postal questionnaire, including the Youth Self-Report (YSR). At age 16 years blood samples taken for DNA extraction for 6 266 adolescents attending the clinical examination. All participants and their parents provided consent to use their data and received Institutional Review Board approval by University of Oulu, and the Ethics committee of the Ostrobotnia Hospital district. More information may be found at: <http://www.oulu.fi/nfbc>.

#### NTR

The Netherlands Twin Register (NTR) is a population-based prospective cohort study which includes new-born twins and multiples from the Netherlands. Recruitment started with birth year 1986. NTR data collection has a focus on growth, development, emotional and behavioural problems and health. Phenotype data on Internalising problems were collected by surveys, in which parents and teachers were asked to rate their offspring / pupils’ behaviour using standardized instruments (Bartels et al., 2007; Ligthart et al., 2019; Van Beijsterveldt et al., 2013). At age 14 years and after, twins and their siblings were asked for self-assessments (Bartels, van de Aa, van Beijsterveldt, Middeldorp, & Boomsma, 2011). Buccal cells and blood for DNA isolation were collected in multiple sub-projects (Scheet et al., 2012). More information may be found at: <https://tweelingenregister.vu.nl/>. The study was approved by the Central Ethics Committee on Research Involving Human Subjects of the VU University Medical Centre, Amsterdam, an Institutional Review Board certified by the U.S. Office of Human Research Protections (IRB number IRB00002991 under Federal-wide Assurance- FWA00017598; IRB/institute codes, NTR 03-180).

#### PNC

The Philadelphia Neurodevelopmental Cohort (PNC) is a large sample of approximately 10,000 genotyped children, adolescents, and young adults with diverse data on a myriad of clinical and cognitive phenotypes (Satterthwaite et al., 2016). Per the original investigators requests, we refer the reader to their previous publications for additional information on the collection of genotypic (Glessner et al., 2010) and clinical data (Calkins et al., 2015; Calkins et al., 2014). The relevant genetic and phenotypic data were downloaded via the Database for Genotypes and Phenotypes (dbGaP Study Accession: phs000607.v3.p2).

#### The Raine Study

The Raine Study is a prospective pregnancy cohort where 2900 mothers where recruited between 1989 and 1991 (Newnham, Evans, Michael, Stanley, & Landau, 1993; Straker et al., 2017). Recruitment took place at Western Australia’s major perinatal centre, King Edward Memorial Hospital, and nearby private practices. Women who had sufficient English language skills, an expectation to deliver at King Edward Memorial Hospital, and an intention to reside in Western Australia to allow for future follow-up of their child were eligible for the study. The Raine Study is known to be one of the largest successfully prospective cohorts richly phenotyped at multiple time points over pregnancy, infancy, childhood adolescence, and young adult. The mothers completed questionnaires regarding their children and the children had physical examinations at ages 1, 2, 3, 6, 8, 10, 14, 17, 20 and 22 years.

#### TEDS

The Twins Early Development Study (TEDS) is a longitudinal twin study that recruited over 16 000 twin pairs born between 1994 and 1996 in England and Wales through national birth records (Haworth, Davis, & Plomin, 2013). More than 10 000 of these families are still involved in study. TEDS was and still is a representative sample of the population in England and Wales. Rich cognitive and behavioural data have been collected from the twins from infancy to emerging adulthood with data collection at ages 2, 3, 4, 7, 8, 9, 10, 12, 14, 16, 18, 19 and 21, enabling longitudinal genetically sensitive study designs. Data have been collected from twins themselves (including extensive web-based cognitive testing), from their parents and teachers, and from the UK National Pupil Database. Genotyped DNA data are available for 10 346 individuals (who are unrelated except for 3 320 dizygotic co-twins). TEDS data have contributed to over 400 scientific papers involving more than 140 researchers in 50 research institutions.

#### TRAILS

Tracking Adolescents’ Individual Lives Survey (TRAILS) is a large prospective population study of Dutch adolescents with bi- or triennial measurements from age 11 years onwards. TRAILS participants were selected from five municipalities in the Northern part of the Netherlands (de Winter et al., 2005). The cohort’s characteristics and database are described in detail elsewhere (Oldehinkel et al., 2015) and at <http://www/trails.nl/>. DNA was extracted from blood samples or (in a few cases) buccal swaps, collected at about age 16. In total, 1,491 children of white European descent were genotyped. The study was approved by the Dutch Central Committee on Research Involving Human subjects (CCMO), and all measurements were carried out with participants’ adequate understanding and written consent.

#### VTSABD

The Virginia Commonwealth University (VCU) arm of the NIDA-funded Gene-Environment-Development Initiative (GEDI) combined existing phenotypic and environmental data from the Virginia Twin Study of Adolescent Behavioral Development (VTSABD) study, a population-based multi-wave, cohort-sequential twin study of adolescent psychopathology and its risk factors, with genome-wide genotyping, generating a genotyped sample of ~900 subjects (Costello et al., 2013; Eaves et al., 1997; Hewitt et al., 1997; Maes, Silberg, Neale, & Eaves, 2007; Simonoff et al., 1997).

#### YFS

The Young Finns study (YFS) is an on-going longitudinal population-based cohort study that includes 3,596 healthy Finnish children and adolescents from six birth cohorts (aged 3, 6, 9, 12, 15, and 18 years at the study baseline in 1980) (Juonala, Viikari, & Raitakari, 2013; Raitakari et al., 2008). The participants have been followed up over 40 years to investigate childhood risk factors for cardiometabolic outcomes in adulthood. The study has been approved by the ethical committee of the Hospital District of Southwest Finland on 20.06.2017 (ETMK:68/1801/2017), and all subjects have given an informed written consent. Data protection will be handled according to current regulations.

### **Supplementary Methods**

#### Deviations from the re-registration

This project was pre-registered at the open science framework on September 12, 2019 ([https://osf.io/edas6](https://osf.io/edas6/)). The N-weighted GWAMA was performed as planned, but there was not enough power in the dataset to perform meta-analyses using genomicSEM. In the initial follow-up analyses, we were unable to estimate genetic correlations between stratified assessments of internalising symptoms, aside from the correlation between overall internalising symptoms and self-reported internalising symptoms, as the z-score of the SNP heritabilities were less than 4, indicating that there was insufficient power to compute these genetic correlations. Finally, as no genome-wide significant variants were identified, we only performed preliminary functional analyses.

#### Sample description

Eligible cohorts received a standard operation protocol detailing the analysis plan. All participating cohorts were population-based cohorts, and many were birth or pregnancy cohorts with long-term follow-up. In general, cohorts started with parental report during childhood and often switched to self-report in adolescence. To maximise the utility of the wealth of information available, cohorts were requested to contribute multiple GWASs for internalising symptoms, for every combination of rater, instrument, and age. Cohorts contributed between 1 to 18 GWASs, with an average of ~5 GWASs per cohort. For more information on the cohorts see Supplementary Table 1-4.

#### Genotyping, quality control, and imputation

Genotyping was performed within each cohort on common genotyping arrays. Cohort-specific pre-imputation quality control was performed, with variant and sample filtering, based on call rate, minor allele frequency (MAF), Hardy-Weinberg equilibrium (HWE), and excessive heterozygosity (see Supplementary Table 5). Cohorts removed samples with non-European ancestry or mismatched sex. The most commonly used genotyping array across cohorts were the Illumina 660K, Illumina 670K, and Affymetrix 6.0 arrays. Cohorts were asked to use genotypes imputed to the 1000 Genomes (1000G) reference set, mapped to build 37 of the human Genome Reference Consortium assembly (GRCh37). 17 cohorts (77.3%) used 1000G phase 3 version 5 as reference set for the imputation, and the remaining 5 (22.7%) imputed to 1000G phase 1 version 3.

#### Univariate GWASs

For all cohorts, local analysts performed univariate GWASs where internalising symptoms score was regressed on the SNP with sex, age, and first five ancestry-based principal components as covariates, and, if necessary, cohort-specific covariates (see Supplementary Table 6). GWASs included autosomal SNPs.

Genome-wide association analyses were stratified by rater, age, and instrument, with a minimum of 450 observations in each stratum. Descriptive statistics for each uploaded GWAS are shown in Supplementary Table 4. Each cohort supplied the phenotypic correlations between the internalising symptom measures within the different strata and the degree of sample overlap between strata. These statistics allowed us to account for dependence due to repeated measurements.

#### Pre-GWAMA QC

Each uploaded file underwent QC using the EasyQC software package (Winkler et al., 2014). SNPs with a genotyping rate below 95% were removed. Variant QC filters on MAF and HWE *p*-value were applied based on the sample size of the GWAS. An info score cut-off of 0.6 and 0.7 was applied to SNPs that were imputed using MACH and IMPUTE, respectively. Reported allele frequencies were compared with an imputation-matched reference file and variants with an absolute difference larger than 0.2 were removed. Supplementary Table 7 reports the number of remaining SNPs before and after QC.
